# Brain Perivascular Space Morphometry Reflects Neuropsychological and Shared Familial Microvascular Architecture Beyond Vascular Risk

**DOI:** 10.64898/2026.01.22.26344665

**Authors:** Alexandra Morozova, Maria del C. Valdés Hernández, Roberto Duarte Coello, Heather Whalley, Andrew McIntosh, Anca-Larisa Sandu Giuraniuc, Gordon D. Waiter, Christopher J McNeil, Douglas Steele, Jennifer A. Macfarlane, Alison Murray, Joanna M. Wardlaw

**Affiliations:** Centre for Clinical Brain Sciences, the University of Edinburgh, Chancellor’s Building, 49 Little France Crescent, Edinburgh EH16 4SB, UK; Department of Anatomy, Third Faculty of Medicine, Charles University, Ruska 87, Prague, 10000, Czechia; UK Dementia Research Institute Centre at the University of Edinburgh, Chancellor’s Building, 49 Little France Crescent, Edinburgh EH16 4SB, UK; Scottish Imaging Network: A Platform for Scientific Excellence (SINAPSE,www.sinapse.ac.uk); University of Aberdeen, Lilian Sutton Building, Foresterhill, Aberdeen AB25 2ZD, UK; University of Dundee, Ninewells Hospital, Dundee DD1 9SY, UK; NHS Tayside, Dundee, DD1 9SY, UK

**Keywords:** perivascular spaces, cerebral microvasculature, MRI, morphometry, familial clustering, neuropsychiatric factors

## Abstract

**Background:** Brain perivascular spaces (PVS) are emerging MRI markers of microvascular function and waste metabolite clearance. While PVS enlargement has been linked to aging and vascular risk, it remains unclear whether PVS morphology reflects shared familial microvascular characteristics and how these are shaped by individual vascular, physiological, and neuropsychiatric factors. In this study, we investigated whether PVS morphometry captures these familial characteristics, further modulated by individual determinants.

**Methods:** We analyzed 1,183 participants from the Stratifying Depression and Resilience Longitudinally (STRADL) family-based cohort, including 324 individuals with first-degree relatives. Automated MRI segmentation quantified PVS volume, count, density, and median length in the centrum semiovale and basal ganglia. Linear mixed-effects models assessed associations with age, hypertension, hair cortisol, depressive symptom scores, and hand grip strength while accounting for familial clustering.

**Results:** In 1050 individuals (59.5% female; mean age 59.3 ±10.1 years) PVS burden increased with age (PVSvolume%ROI β=0.18, 95%CI[0.11, 0.26], p<0.0001), current depressive symptoms across both regions (PVS density: CSO, β=0.092, [0.023, 0.16], p=0.009; BG, β=0.11, [0.043, 0.18], p=0.002), and with higher hair cortisol (PVS count β=0.08, [0.003, 0.15], p=0.041) and weaker grip strength (PVSvolume%ROI β=-0.09, [-0.16, -0.02], p=0.013), in the centrum semiovale. Familial clustering was significant for PVS volume (β=0.22, [0.096, 0.52], p=0.013) and median length (β=0.28, [0.16, 0.49], p=0.0003), independent of other factors.

**Conclusions:** PVS morphology reflects neuropsychiatric and shared familial microvascular architecture, with both inherited and individual factors contributing to PVS burden and morphometry, and supporting the use of PVS morphometry as a neuroimaging marker of cerebral microvascular health.

## Introduction

Brain perivascular spaces (PVS) are MRI-visible compartments surrounding cerebral perforating vessels, serving as conduits for waste clearance and maintaining microvascular and brain homeostasis [1,2,3]. Enlargement of PVS has been associated with aging, hypertension, and neurovascular and neurodegenerative conditions, including cerebral small vessel disease,multiple sclerosis, and dementia[4,5,6,7]. PVS morphology exhibits high inter-individual variability [2], with genetics being a substantial predisposing factor [8,9], as demonstrated by recent large genome-wide association studies [10,11]. However, genetic association alone does not fully capture the extent to which microvascular and perivascular structure is shared within families, where inherited factors coexist with shared developmental and environmental influences.

Microvascular function may be modulated by neuropsychiatric and physiological factors, including chronic stress, depressive symptoms, and physical strength, through overlapping and potentially interacting pathways such as inflammation, oxidative stress, and vascular dysregulation [12,13–15]. Chronic stress is a well-established pro-inflammatory trigger that disrupts microcirculatory function [12] and a risk factor for cardiovascular and neurodegenerative disorders [16], while depressive symptoms have been linked to neuroinflammation and decreased cerebral blood flow [16,17]. Reduced muscular strength is associated with increased cardiometabolic risk, cardiovascular and overall mortality, and cognitive decline [18]. These factors may act in concert to influence small vessel integrity, with PVS morphology reflecting both individual determinants and familial impact.

To date, few studies have examined the relationship between neuropsychiatric factors and PVS. Existing work suggests associations between stress- or trauma-related exposures and increased PVS burden [19], potentially mediated by neuroinflammatory processes [20], as well as links between PVS burden and post-stroke depression [21]. However, the extent to which neuropsychiatric factors independently influence PVS enlargement and morphometric features remains unclear. Moreover, while the heritability of PVS burden has been demonstrated [10,11], whether, and the extent to which, PVS morphometry reflects shared familial microvascular architecture has not been established.

In this study, we examined familial and individual determinants of PVS burden and morphology in a large, population-based cohort, including familial clustering of PVS morphometric features among first-degree relatives to distinguish shared familial from individual influences, while evaluating associations with neuropsychiatric factors (chronic stress assessed via hair cortisol and depressive symptoms measured by the Quick Inventory of Depressive Symptomatology [22]) and physical strength (hand grip [23]). Moving beyond conventional visual ratings or single quantitative metrics, we applied a comprehensive computational morphometric approach, quantifying PVS volume, count, density, and median length to capture distinct dimensions of PVS architecture. White matter hyperintensities, established markers of small vessel disease [24] topologically linked with PVS [25], were included as covariates to isolate effects specific to PVS. We hypothesised that familial membership would confer similarity in PVS morphometry, and that age, vascular risk, higher stress, and depressive symptoms would associate with greater PVS burden, whereas stronger hand grip would associate with lower PVS burden as evidence of microvascular health.

## Methods

### Sample

The dataset comes from the Stratifying Depression and Resilience Longitudinally (STRADL) study [26], a population-based longitudinal study examining the biological and psychological mechanisms of depression and resilience. The flow chart of the recruitment and final sample used in this study is shown in flow chart, **Supplementary Figure 1**. Briefly, from the 21,525 members of the Generation Scotland Scottish Family Health Study (GS:SFHS) cohort [32] eligible for re-contact, 5,649 were invited to participate in STRADL. From them, 646 declined and 3,358 did not respond. From those who consented to participate, we considered for this study 1,183 subjects that had all relevant clinical and demographic data available, with MRI assessment performed at two locations, Aberdeen (51%) and Dundee (49%), in Scotland, United Kingdom, using 3T magnetic resonance imaging scanners. From them, 859 individuals were unrelated, and 324 were part of 136 families of between two and six members – all first-degree relatives. It is worth noting that 128 had missing MRI data (i.e., either incomplete MRI scan or missing or corrupted MRI sequences). Therefore, we used MRI data from 1,055 study participants.

#### Image processing

The neuroimaging protocol of the primary study that provided data for this analysis is published [26]. We used previously defaced T1-weighted, T2-weighted and fluid attenuated inversion recovery (FLAIR) magnetic resonance images (MRI), as well as the brain parcellation done by Freesurfer v 6.0 (https://surfer.nmr.mgh.harvard.edu/), all provided by the Generation Scotland Data Access Committee (https://genscot.ed.ac.uk/for-researchers/access), together with the rest of the variables used in the analyses. We co-registered all MRI sequences to the nearly-isotropic (1 x 0.94 x 0.94 mm^3^) T1-weighted space, where the Freesurfer segmentations are, and obtained the intracranial volume using FLIRT [27] and BET2 [28]. We applied Gaussian clustering to the multisequence (4D) volume obtained by concatenating the three co-registered MRI sequences to extract the brain tissue separately from the pure CSF, the meningeal layers and main venous pathways. We obtained initial priors of the basal ganglia (BG) and white matter regions (mainly centrum semiovale (CSO), excluding the internal and external capsules that were included in the BG region) from Freesurfer segmentations. We refined them to ensure that venous pathways, membranes and CSF were not included, by applying the masks generated from the multispectral Gaussian clustering segmentations. Then, we mapped our (3D) regions of interest (ROIs) to the T2-weighted space, of higher spatial resolution (i.e., 0.5 x 0.48 x 0.48 mm^3^), where we performed the PVS segmentations using the thresholded output from the Frangi filter as described in Ballerini et al. [29]. From the PVS segmentations we calculated the percentage volume and the density of PVS in each ROI (PVS counts in a ROI volume), and determined the average PVS length per ROI using the Bezier curves approximation [30]. WMH were segmented automatically using nn-UNet, a self-configuring U-shaped neural network for segmenting biomedical images using deep-learning [31], previously trained on a set of 162 FLAIR MRI data from individuals from studies of ageing and patients with mild stroke, all with different degrees of small vessel disease [32]. The out-of-the-box nn-UNet pre-trained model used is 3D with batch size of two and patch size of 112 x 160 x 128 voxels. It uses as input FLAIR images, resampled to 0.94 x 0.94 x 0.90 mm^3^ voxel-size resolution.

### Overview of methods for glucocorticoid extraction / grip strength / QIDS/ hypertension

Demographic data (age and sex), were obtained within the GS:SFHS [33] between 2006 and 2011. Grip strength, hair sample, blood pressure, and QIDS were obtained at the same time as the MRI in an in-person assessment wave in 2019 for the STRADL study (See **Supplementary Figure 1**). Grip strength was determined using a Patterson Medical Jamar hand dynamometer. A small sample (i.e., approximately 3mm diameter) of hair was collected from the posterior vertex region of the head for extracting cumulative cortisol using organic solvents in pulverised hair and processed using liquid chromatography-mass spectroscopy [26].

### Statistical Analysis

We applied a linear mixed effects model (LME) to account for possible correlations in the PVS characteristics among first-degree relatives. Family ID was included as a random intercept to model shared variance within families. Age, hypertension, hair cortisol, grip strength, WMH burden and QIDS score were entered as fixed effects to examine their influence on the PVS volume. This model enabled to assess both within-family and between-family variation, including comparisons between family members and unrelated individuals, providing insight into the contribution of familial factors on the outcome. Due to skewed distributions, WMH burden and hair cortisol concentration were log-transformed before inclusion in the models.

Since the dataset contained both individuals and families, a sensitivity analysis was performed by repeating the analysis on a sample that excluded individuals without family members (i.e., restricting the sample to families with at least two members). Family ID was added as a random intercept, as in the primary model, while age was added as a random covariate to assess potential differences in its effect across families. We did not exclude participants with a history of stroke or any other incidental finding in their brain scans.

All statistical analyses were performed using the Stata software package (version 18, Stata Corp. 2023. *Stata Statistical Software: Release 18*. College Station, TX: Stata Corp LLC.). No correction for multiple comparisons was applied.

## Results

### Descriptive Statistics

The derivation of the study sample (n = 1050) from the initial cohort (n = 1183) is shown in Supplementary Figure 1, with exclusions largely due to unavailable MRI data (n = 103) or missing MRI sequences required for analysis (n = 25). Demographic, clinical, and neuroimaging characteristics are presented in **Table 1**. No statistically significant differences in the average parameters evaluated were observed between the sample included and the total sample comprised of those who consented and underwent the relevant clinical tests for this study (see values in **Table 1**). The distribution of family sizes is summarized in **Table 2**. Each family size represents the number of 1^st^ degree members per family. The mean age was 59.3+/-10.1 years, and 625/1050 (59.5%) were female. Most did not have hypertension or prior stroke.

**Table 1.**
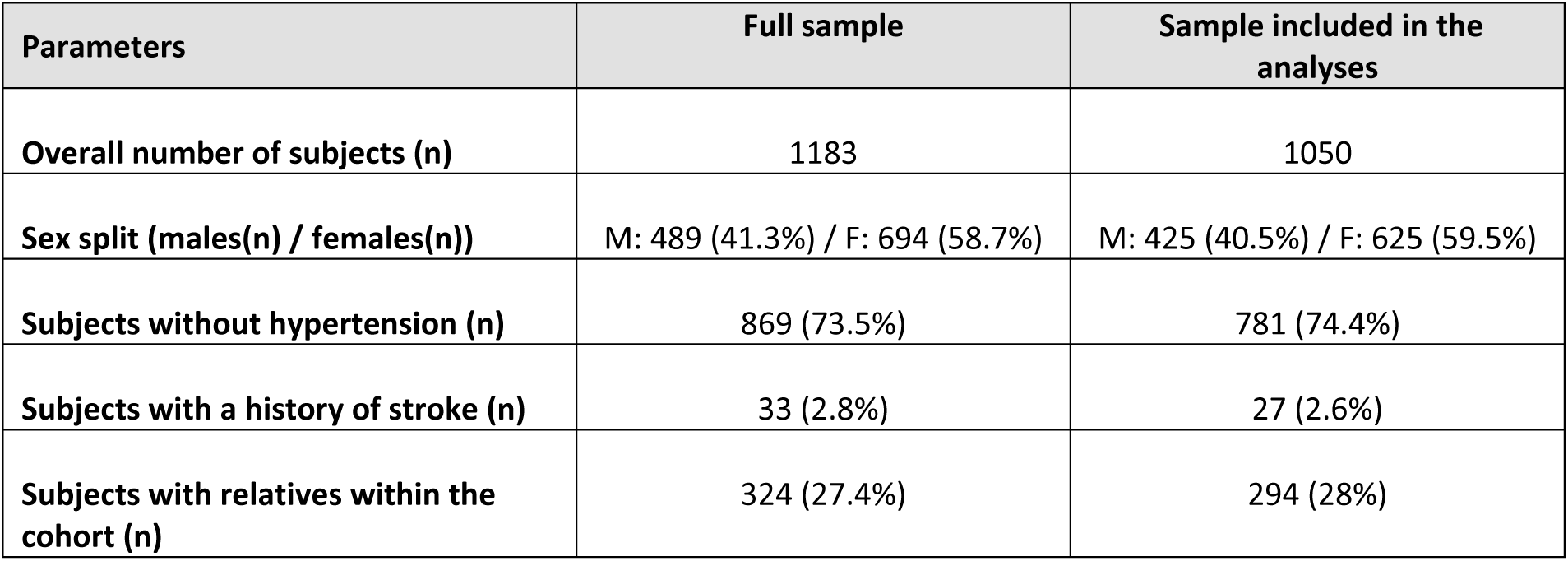
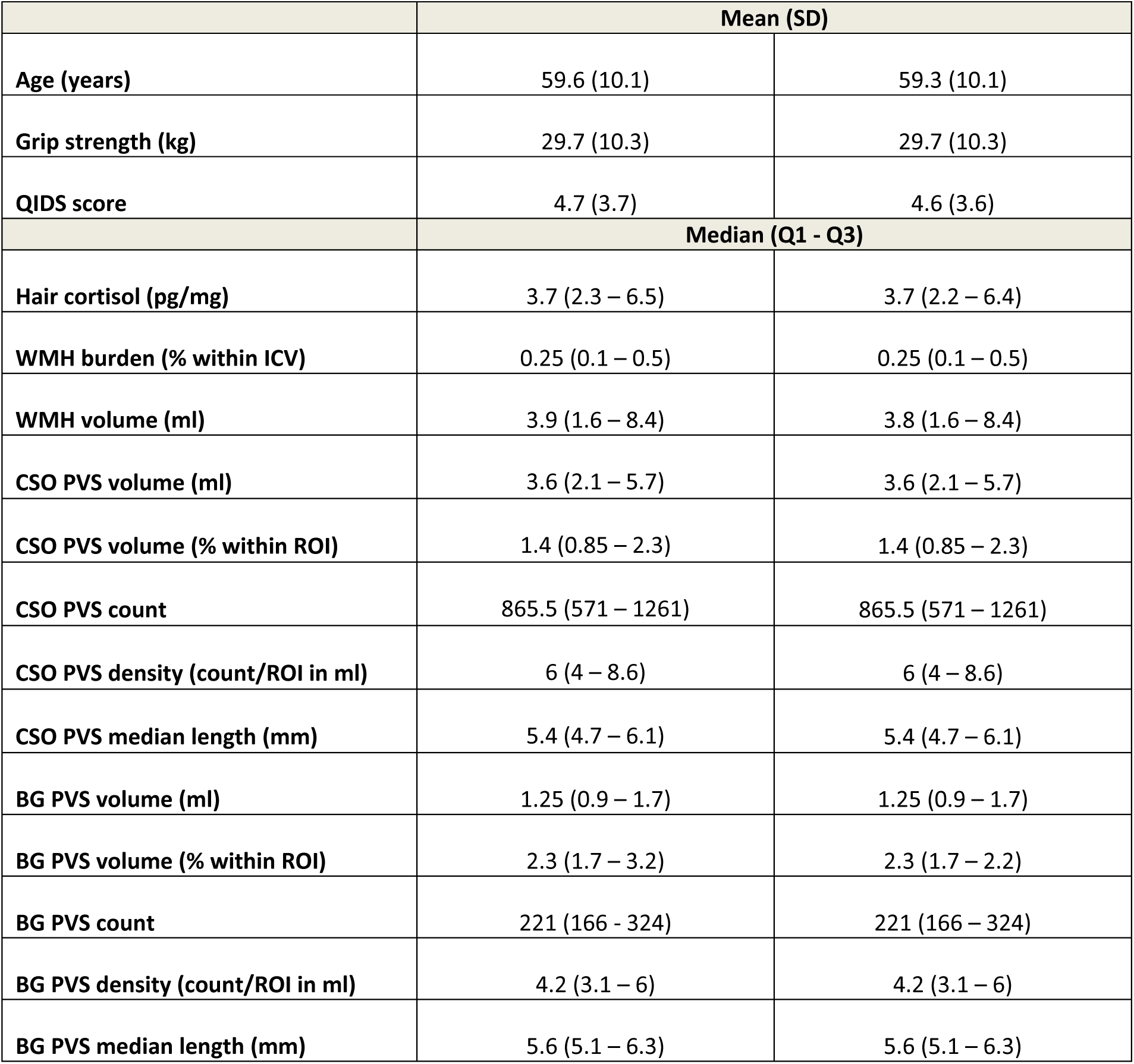
Study data overview

**Table 2.** Distribution of Family Cluster Sizes in the Sample (First-Degree Relatives Only).

| Family Size | Number of participants in the full sample | Percentage | Number of participants in the analysis | Percentage |
| --- | --- | --- | --- | --- |
|  | (n=1183) |  | (n=1050) |  |
| 1 | 859 | 72.6% | 756 | 72% |
| 2 | 204 | 17.2% | 185 | 17.6% |
| 3 | 63 | 5.3% | 57 | 5.4% |
| 4 | 36 | 3% | 33 | 3.1% |
| 5 | 15 | 1.3% | 14 | 1.3% |
| 6 | 6 | 0.5% | 5 | 0.5% |

### Primary Analyses

A Linear Mixed Effects (LME) model was built for each PVS measurement (PVS volume % in the regions of interest (ROI), count, density, and median length) as an outcome in both the centrum semiovale (CSO) and the basal ganglia (BG). The results from all models are summarised in **Table 3** (standardised values) and in the **Supplementary Table** (non-standardised values). The stated β-coefficients are standardised with 95% Confidence Intervals in square brackets unless otherwise stated. Associations between PVS volume (% in ROIs; the PVS characteristic representative of the PVS burden) and statistically significant covariates are shown in **Figures 1 and 2**.

**Figure 1.**
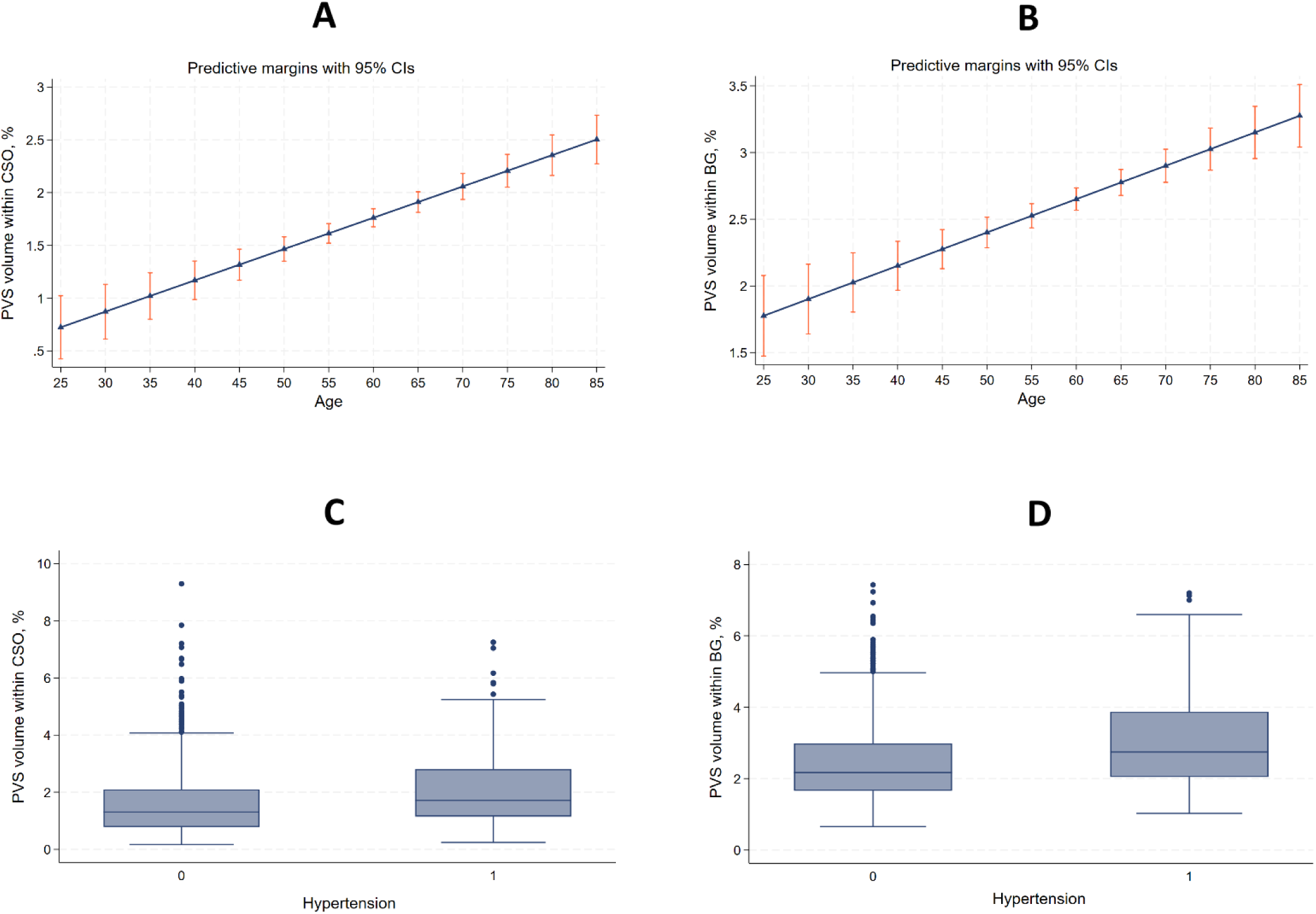
Association between age (in years) and PVS volume (as % of the region of interest) in the centrum semiovale (A), and basal ganglia (B), and association between hypertension (yes=1 and no=0) and PVS volume (as %in the region of interest) in the centrum semiovale (C), and basal ganglia (D).

**Figure 2.**
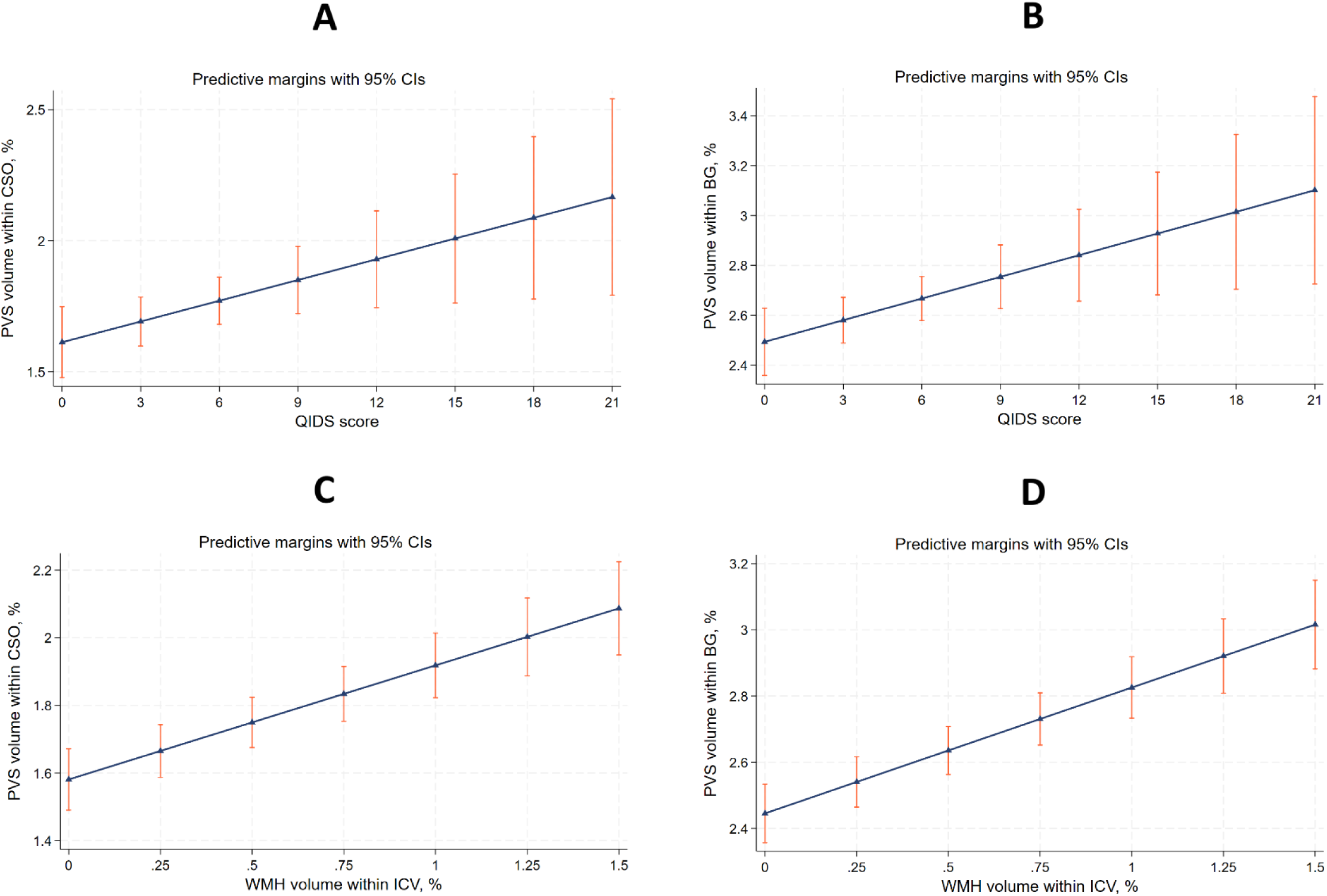
Association between the QIDS score and PVS volume (as % of the region of interest) in the centrum semiovale (A), and basal ganglia (B), and association between WMH volume within ICV in % and PVS volume (as % in the region of interest) in the centrum semiovale (C), and basal ganglia (D). WMH volume was truncated at the 95th percentile for visualization purposes only.

**Table 3.** Linear mixed effects models - standardized results (β, standard error (SE), p-values, 95% CI)

|  | *Centrum Semiovale* |  |  |  | *Basal Ganglia* |  |  |  |
| --- | --- | --- | --- | --- | --- | --- | --- | --- |
|  | PVS volume (% in ROI) | PVS count | PVS density (count per mm3) | PVS median length (mm) | PVS volume (% in ROI) | PVS count | PVS density (count per mm3) | PVS median length (mm) |
| *Fixed Effect* |  |  |  |  |  |  |  |  |
| **Intercept** | -0.039 (0.04) | -0.02 (0.042) | -0.023 (0.042) | -0.033 (0.04) | -0.07 (0.041) | -0.043 (0.043) | -0.045 (0.042) | -0.065 (0.041) |
| **Age (years)** | **0.18 (0.038),**<br>**p=0.000,**<br>**[0.11 0.26]** | **0.12 (0.042),**<br>**p=0.005,**<br>**[0.036 0.2]** | **0.16 (0.041),**<br>**p=0.000,**<br>**[0.076 0.24]** | **0.15 (0.039),**<br>**p=0.000,**<br>**[0.074 0.23]** | **0.17 (0.04),**<br>**p=0.000,**<br>**[0.088 0.24]** | 0.03 (0.042),<br>p=0.478,<br>[-0.053 0.11] | **0.092 (0.042),**<br>**p=0.027,**<br>**[0.01 0.17]** | **0.10(0.04),**<br>**p=0.011,**<br>**[0.023 0.18]** |
| **Hypertension (yes/no)** | 0.15 (0.08),<br>p=0.067,<br>[-0.01 0.31] | 0.13 (0.087),<br>p=0.14,<br>[-0.042 0.30] | 0.16 (0.085),<br>p=0.068,<br>[-0.011 0.32] | 0.6 (0.83),<br>p=0.47,<br>[-0.10 0.22] | **0.32 (0.084),**<br>**p=0.000,**<br>**[0.16 0.49]** | **0.22 (0.088),**<br>**p=0.014,**<br>**[0.044 0.39]** | **0.23 (0.087),**<br>**p=0.008,**<br>**[0.06 0.40]** | **0.25 (0.084),**<br>**p=0.002,**<br>**[0.09 0.42]** |
| **Hair cortisol (log)** | 0.06 (0.034),<br>p=0.079,<br>[-0.007 0.13] | **0.08 (0.037),**<br>**p=0.041,**<br>**[0.003 0.15]** | **0.074 (0.036),**<br>**p=0.041,**<br>**[0.003 0.14]** | 0.015 (0.035),<br>p=0.65,<br>[-0.053 0.084] | -0.0013 (0.036),<br>p=0.97,<br>[-0.071 0.069] | 0.1(0.038),<br>p=0.79,<br>[-0.064 0.084] | 0.015 (0.037),<br>p=0.69,<br>[-0.058 0.088] | -0.059 (0.036),<br>p=0.098,<br>[-0.13 0.011] |
| **Grip strength (kg)** | **-0.09 (0.04),**<br>**p=0.013,**<br>**[-0.16 -0.02]** | 0.011 (0.039),<br>p=0.77,<br>[-0.066 0.088] | **-0.08 (0.039),**<br>**p=0.038,**<br>**[-0.16 -0.0045]** | -0.045 (0.037),<br>p=0.226,<br>[-0.12 0.028] | -0.047 (0.038),<br>p=0.21,<br>[-0.12 0.027] | 0.026 (0.04),<br>p=0.51,<br>[-0.052 0.10] | -0.047 (0.040),<br>p=0.23,<br>[-0.12 0.30] | -0.011 (0.038),<br>p=0.764,<br>[-0.085 0.063] |
| **QIDS score** | **0.074 (0.033),**<br>**p=0.026,**<br>**[0.009 0.14]** | **0.079 (0.036),**<br>**p=0.027,**<br>**[0.009 0.15]** | **0.092 (0.035),**<br>**p=0.009,**<br>**[0.023 0.16]** | -0.043 (0.034),<br>p=0.207,<br>[-0.11 0.024] | **0.085 (0.034),**<br>**p=0.014,**<br>**[0.017 0.15]** | **0.10 (0.036),**<br>**p=0.004,**<br>**[0.032 0.17]** | **0.11 (0.036),**<br>**p=0.002,**<br>**[0.043 0.18]** | -0.050 (0.034),<br>p=0.147,<br>[-0.12 0.018] |
| **WMH burden**<br>**within ICV**<br>**(log)** | **0.13 (0.04),**<br>**p=0.002,**<br>**[0.048 0.21]** | **0.13 (0.044),**<br>**p=0.003,**<br>**[0.044 0.22]** | **0.11 (0.043),**<br>**p=0.009,**<br>**[0.028 0.2]** | **0.1 (0.042),**<br>**p=0.014,**<br>**[0.021 0.18]** | **0.093 (0.042),**<br>**p=0.028,**<br>**[0.01 0.18]** | 0.035 (0.044),<br>p=0.436,<br>[-0.0520.12] | 0.032 (0.044),<br>p=0.46,<br>[-0.054 0.12] | **0.15 (0.042),**<br>**p=0.000,**<br>**[0.065 0.23]** |
| *Random Effect* |  |  |  |  |  |  |  |  |
| **Family ID** | **0.22 (0.10),**<br>**p=0.013,**<br>**[0.096 0.52]** | 0.081 (0.12),<br>p=0.24,<br>[0.005 1.33] | 0.11 (0.12),<br>p=0.17,<br>[0.014 0.89] | **0.28 (0.079),**<br>**p=0.0003,**<br>**[0.16 0.49]** | 0.057 (0.11),<br>p=0.304,<br>[0.0013 2.55] | m.o. | 7.55E-6 (0.005),<br>p=1.0,<br>[0 0] | 0.094 (0.093),<br>p=0.147,<br>[0.013 0.65] |

Legend: PVS: MRI-visible enlarged perivascular spaces, ROI: Region Of Interest, m.o.: model overfitting

The variance component for Family ID was significant in the CSO with the percentage of PVS volume in this region as the outcome (standardized β=0.22, 95%CI [0.096, 0.52], p=0.013), and PVS median length (β=0.28, [0.16, 0.49], p=0.0003), indicating that a substantial portion of the variance in CSO is explained by familial factors. In the BG, Family ID effect was not associated with PVS volume%ROI.

Age was a highly significant predictor of the PVS burden in the CSO across all models for PVS volume%ROI (β=0.18, [0.11, 0.26], p<0.0001), density (β=0.16, [0.076, 0.24], p<0.0001), median length (β=0.15, [0.074, 0.23], p<0.0001), and count (β=0.12, [0.036, 0.2], p=0.005), **Figure 1A**, **Table 3** **and Supplementary Table**. In the BG, age was associated with PVS volume%ROI (β=0.17 [0.088, 0.24], p<0.001), **Figure 1B**, PVS density (β=0.092, [0.01, 0.17], p=0.027), and PVS median length (β=0.1, [0.023, 0.18], p=0.011) (see **Table 3**).

Hypertension was not associated with PVS measurements in the CSO apart from a trend for PVS volume%ROI (β=0.15, [-0.01, 0.31], p=0.067, **Figure 1C**), but was associated with all PVS measures in the BG: PVS volume%ROI (β=0.32, [0.16, 0.49], p<0.001, **Figure 1D**); PVS count (β=0.22, [0.044, 0.39],p=0.014); PVS density (β=0.23, [0.06, 0.40],p=0.008); and PVS median length (β=0.25, [0.09, 0.42], p=0.002), **Table 3**).

The associations of the QIDS depression score with PVS were the same in the CSO and the BG: PVS volume%ROI (CSO, β=0.07, [0.009, 0.14], p=0.026), **Figure 2A**; BG, β=0.085, [0.017, 0.15], **Figure 2B**); PVS count (CSO, β=0.079, [0.009, 0.15], p=0.027, BG, β=0.1, [0.032, 0.17], p=0.004); PVS density (CSO, β=0.092, [0.023, 0.16], p=0.009; BG, β=0.11, [0.043, 0.18], p=0.002). There were no significant associations between the QIDS score and PVS median length in either of the brain regions.

WMH volume (% within ICV) was associated with PVS burden in the CSO across all models: PVS volume%ROI (β=0.13, [0.048, 0.21], p=0.002, **Figure 2C**), PVS count (β=0.13, [0.044, 0.22], p=0.003), PVS density (β = 0.11, [0.028, 0.2], p=0.009), and PVS median length (β = 0.1, [0.021, 0.18], p= 0.014). In the BG, WMH volume (% within ICV) was associated with the PVS volume%ROI (β= 0.093, [0.01, 0.18], p=0.028, **Figure 2D**) and PVS median length (β=0.15, [0.065, 0.23], p<0.001).

Grip strength was negatively associated with increased percentage of PVS volume%ROI (β=-0.09, [-0.16, -0.02], p=0.013, **Figure 3A**) and PVS density (β=-0.08, [-0.16, -0.0045], p=0.038, **Figure 3B**) in the CSO. It was not associated with any PVS measurements in the BG.

**Figure 3.**
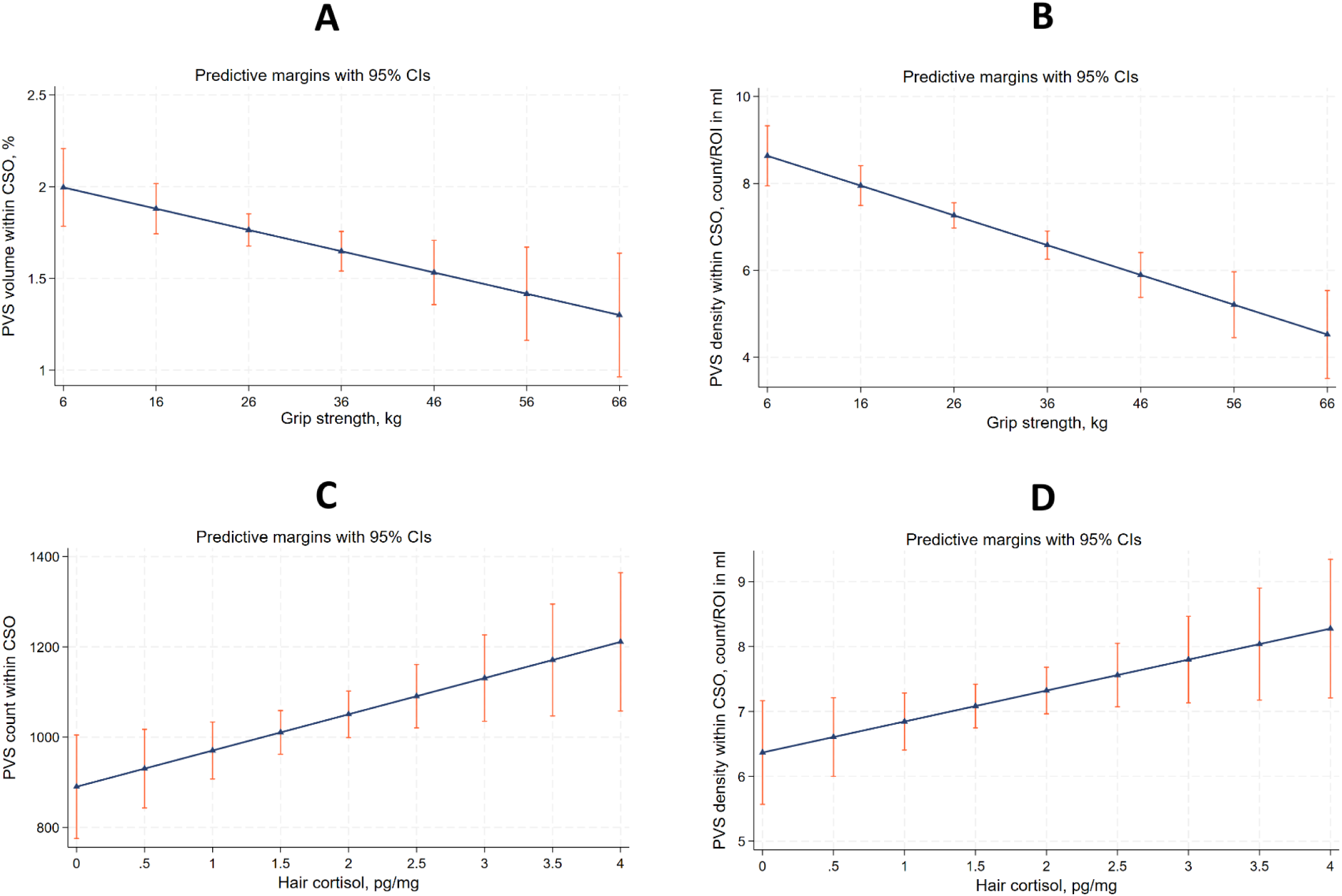
Associations between PVS measures in the centrum semiovale (CSO) and grip strength and hair cortisol. Panels A and B show grip strength (kg) associations with PVS volume (as % of the region of interest) and PVS density (count per region of interest, %). Panels (C–D) show hair cortisol (pg/mg, log-transformed) associations with PVS count and PVS density (count per region of interest, %).

Hair cortisol level was associated with two CSO PVS measurements: PVS count (β=0.08, [0.003, 0.15], p=0.041, **Figure 3C**) and PVS density (β=0.074, [0.003, 0.14], =0.041, **Figure 3D**).

Association with CSO PVS volume (as % in the CSO ROI) was of borderline significance (β=0.06, [-0.007, 0.13], p=0.079). There were no associations between hair cortisol and the BG PVS measurements.

### Sensitivity Analysis

The sensitivity analysis was performed for the two models with the significant random effect of Family ID – the model with the PVS volume%ROIs as outcome, and the model with the PVS median length as outcome, both in the CSO in two parts (**Table 3**). In the first part, we replicated the original model but restricted the sample to families with at least two members, excluding individuals without family members. This allowed us to assess whether being in the same family affected the PVS association (outcome). **Figure 4** shows PVS in the CSO on T2 MRI in three generations of the same family. In the second part, we added age as a random covariate to the model, which revealed that age had substantially different effects across families. The random effect of Family ID remained significant in both parts of the sensitivity analysis for both models for PVS volume%ROI (β=0.22, [0.096, 0.52], p=0.013) and median length (β=0.28, [0.16, 0.49], p=0.0003), and the model fit was very similar (Akaike Information Criterion (AIC) PVS volume%ROI 669.95 vs 667.56; PVS median length AIC 695.7 vs 693.01), with the model including age as a random covariate showing slightly stronger associations.

**Figure 4.**
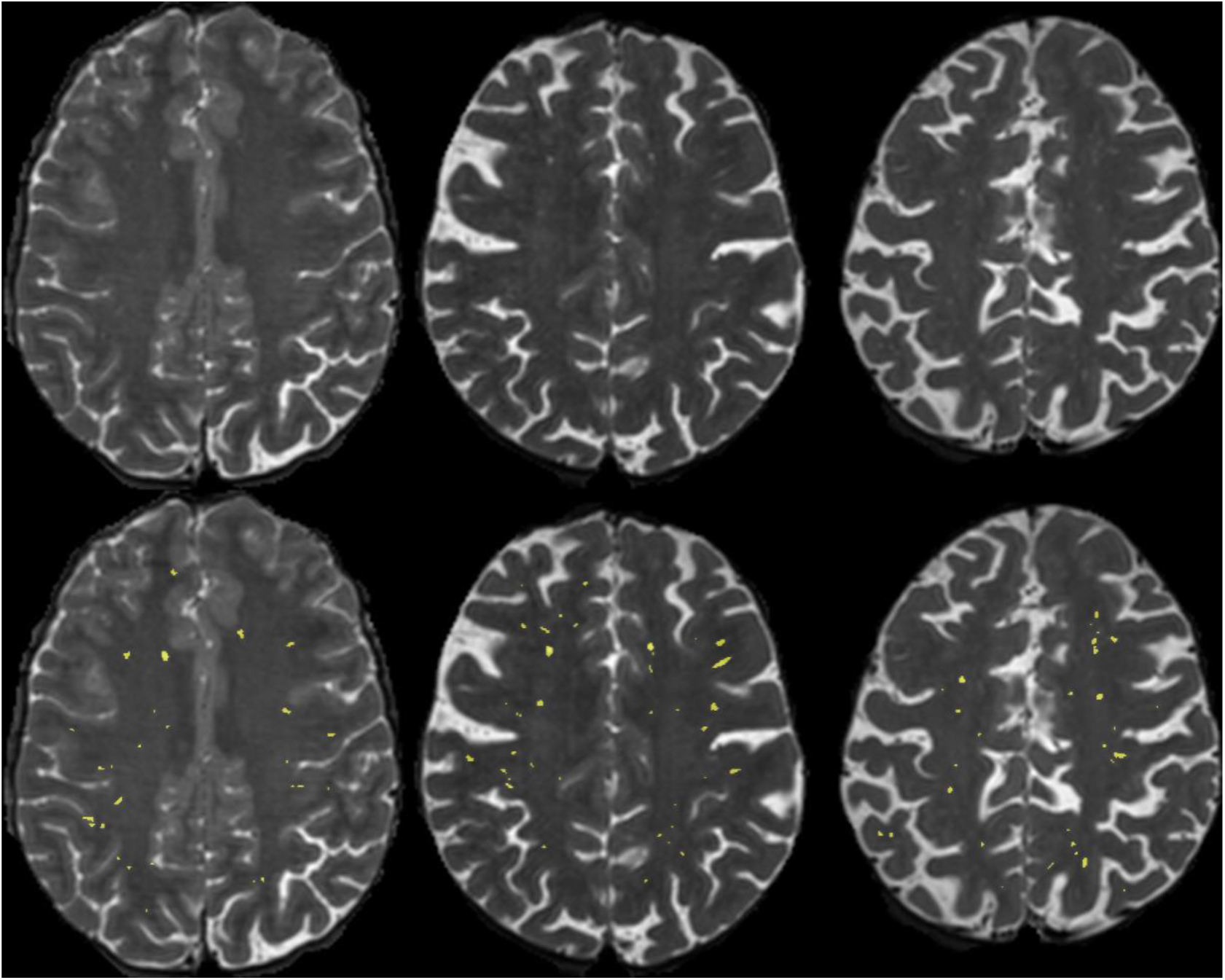
Equivalent axial T2-weighted MRI slice in the centrum semiovale, with the PVS segmented (bottom row) in yellow, in three consecutive generations of the same family (younger to older: left to right). Observe similar number of PVS counted regardless of age differences, and similarities in the topological (i.e., spatial) distribution patterns in the [two pairs of] consecutive generations.

## Discussion

Our study demonstrates that PVS morphology is shaped by familial, vascular, neuropsychiatric, and physiological factors, providing novel insight into cerebral microvascular structure and function. Evidence of familial clustering in CSO PVS volume and median length highlights inherited contributions, while vascular, neuropsychiatric, and physiological determinants further modulate these features. These results underscore the multifactorial determinants of cerebral microvasculature and support PVS morphometry as a sensitive neuroimaging marker of microvascular function.

### Familial influences

Significant familial clustering was observed for CSO PVS volume and median length, independent of individual determinants (**Figure 4**). This finding suggests that perivascular architecture is partially governed by shared genetic and epigenetic influences, consistent with previous heritability studies linking vascular morphology to genetic variation [10,11,34,35]. While age-related perivascular changes are universally observed, their trajectory may vary within families according to other exposures, and between families reflecting inherited susceptibility to microvascular remodeling.

### Vascular factors

Age and hypertension were strongly associated with PVS measures, aligning with previous reports [1, 4]. Age effects were consistent across CSO and BG measures, except for BG PVS count, likely reflecting the lower absolute PVS numbers in this region. Hypertension showed stronger associations in the BG and weaker effects in the CSO. Overall, pathological vascular changes are likely not restricted to the BG but may first be apparent in its small vessels, which perhaps are most proximate to systemic factors like blood pressure. In contrast, the CSO penetrating arterioles are further from the systemic vasculature and have different perivascular membranes so may be less sensitive to systemic effects. The binary nature of our hypertension variable may limit sensitivity, as cumulative blood pressure better captures continuous vascular impact [36].

### Neuropsychiatric factors

Chronic stress, measured via hair cortisol, was associated with increased CSO PVS count and density, but not with BG PVS. Cortisol predominantly affects cortical regions, promoting neuroinflammation and impairing perivascular clearance [19,20,37]. Depressive symptoms, assessed via QIDS at the time of MRI, were associated with higher PVS volume, count, and density in both regions, suggesting that stress and depression may influence perivascular clearance through neuroinflammatory and/or metabolic mechanisms [38,39]. Using continuous measures at the time of MRI likely captured subtle associations beyond those identifiable using categorical yes/no depression diagnoses, given that depression is often under-reported [40].

### Physical strength

Hand grip strength inversely correlated with CSO PVS volume and density, indicating that better physical fitness may preserve microvascular health. Hand grip is a validated marker of cardiovascular, cognitive, and general health [41–43], supporting its potential as a simple proxy for perivascular function. Interestingly, the primary motor and sensory white matter pathways make up a prominent proportion of the CSO as they fan out to the cortex, whereas the same pathways are tightly bunched taking up a smaller proportion of the BG region. Perhaps this is why a PVS-grip strength association was detected in the CSO rather than the BG.

### Regional differences and WMH

WMH burden correlated with PVS measures, consistent with some prior studies [44–47]. Adjusting for WMH allowed isolation of PVS-specific associations, consistent with PVS morphological changes reflecting early microvascular changes preceding overt WMH [48,49]. Regional differences between BG and CSO likely reflect anatomical and functional distinctions, including periarteriolar membranes, perhaps venous drainage [48,49] and compensatory mechanisms, shaping PVS susceptibility to vascular and physiological stress.

Overall, neuropsychiatric and physiological factors may act in concert, alongside vascular influences, to modulate PVS morphology. While familial clustering establishes baseline microvascular architecture, individual determinants such as stress, depressive symptoms, hypertension, and physical strength further modify PVS burden. This emphasizes that both inherited and non-familial factors converge to determine cerebral microvascular structure.

### Limitations

The cross-sectional design precludes causal inference. Familial clustering was assessed in a subset of participants, potentially limiting generalizability. Only 8.8% of participants had a formal depression diagnosis, but continuous QIDS scores at MRI provided robust measures of symptom severity. Longitudinal studies are needed to better characterize PVS dynamics and familial influences over time.

## Conclusion

PVS morphology reflects a shared familial microvascular architecture further modulated by vascular, neuropsychiatric, and physiological factors. These findings highlight inherited and individual contributions, demonstrate regional differences in PVS burden, and support PVS morphometry as a non-invasive neuroimaging marker of cerebral microvascular function.

## Data Availability

All data referred to in the manuscript is available upon request. Details can be found at https://genscot.ed.ac.uk/for-researchers/access

https://genscot.ed.ac.uk/for-researchers

https://shiny.igc.ed.ac.uk/GS_Data_Explorer/

https://doi.org/10.1016/j.cccb.2025.100419

## Acknowledgements

We would like to thank all of the Generation Scotland participants for their contribution to this study. We also thank the research assistants, clinicians and technicians for their help in collecting the data, and Generation Scotland administration for making it available for research. This study is funded by the Hilary and Galen Weston Foundation under the Novel Biomarkers 2019 scheme [ref UB190097l] administered by the Weston Brain Institute, the Fondation Leducq Network of Excellence for the Study of Perivascular Spaces in Small Vessel Disease (16 CVD 05), and the UK Dementia Research Institute [award number UK DRI-4002] through UK DRI Ltd, funded by the UK Medical Research Council, Alzheimer’s Society, and Alzheimer’s Research UK. Generation Scotland received core support from the Chief Scientist Office of the Scottish Government Health Directorates [CZD/16/6] and the Scottish Funding Council [HR03006] and is currently supported by the Wellcome Trust [216767/Z/19/Z]. This study was also supported and funded by the Wellcome Trust Strategic Award ‘Stratifying Resilience and Depression Longitudinally’ (STRADL) [References 104036/Z/14/Z and 220857/Z/20/Z]. We acknowledge the support of the British Heart Foundation [RE/18/5/34216], the Row Fogo Charitable Trust [BRO-D.FID3668413] (MVH, JMW), and Charles University grants to AM: Cooperatio 36 - Medical Diagnostics and Basic Medical Sciences, and Cooperatio 38 – Neurosciences.

